# AI vs. Traditional ultrasound study in Congenital Heart Defect Detection: A Systematic review

**DOI:** 10.1101/2025.04.05.25325217

**Authors:** Shabana Jassim, Shaheen Basheer

## Abstract

**Introduction:** Prenatal detection rates for CHD have increased with improved ultrasound technology and imaging, the use of the first trimester fetal echocardiography, and standardization of the views required in the fetal echocardiogram. Moreover accurate prenatal detection of CHD, particularly complex CHD, is an important contributor to improved survival rates for patients with CHD, however training and availability of expertise is still a limitation in screening. Artificial intelligence has already taken the world by storm, by surpassing human limitations and capabilities. Applications of artificial intelligence in the field of clinical diagnostics holds immense potential. Forty original, primary research studies that studied the role of Artificial Intelligence/ Machine Learning in detecting prenatal congenital heart defects in comparison to fetal echocardiography by traditional ultrasound techniques, were selected and the extracted data was assessed for the accuracy of AI-driven prenatal ultrasound screening for the detection of congenital heart defects.

**Rationale:** Owing to the new technology, lack of generalized access to such information systems and limited availability of advanced AI algorithms in older ultrasound machines at present, well-designed comprehensive research and systematic analyses are still very few and there is much gap in our understanding of this very valuable tool in clinical application and diagnoses.

**Objectives:** To assess the diagnostic accuracy of artificial intelligence algorithms in detecting various congenital heart defects in comparison to traditional ultrasound based fetal echocardiography, across various gestational ages.

**Methods:** A comprehensive search was conducted across databases and registers, identifying 496 records. After removing duplicates and excluding studies based on eligibility criteria, 40 original primary studies were included. Data was extracted on types of congenital heart defects detected, sensitivity of detection, timing of scan, with comparative analysis conducted on detection of the same with traditional ultrasound fetal echocardiography. We searched for academic papers using PubMed, Semantic search tools and journals. Of the papers most relevant to the question that were retrieved, both authors rigorously and independently studied and summarized the abstracts to exclude the studies that did not meet the outlined criteria. Even though some well-planned comprehensive studies were found, they lacked certain key inclusion criteria, and were therefore excluded from the review. 40 studies were shortlisted for the purpose of this study, based on the inclusion criteria. Only studies that involve prenatal ultrasound screening of human fetuses for congenital heart defects, that evaluate AI/machine learning algorithms for detecting congenital heart defects in comparison reporting outcome as at least one quantitative measure of diagnostic accuracy (sensitivity, specificity or accuracy) were included. Study Designs were limited to primary research studies (prospective or retrospective). Gestational age at study was also noted. All Non-human studies/ Duplicate publications/ Systematic or narrative reviews and studies without key information related to the question asked, were excluded.

**Results:** Across gestational ages, artificial intelligence algorithms detect congenital heart defects with 95-96% accuracy, exceeding traditional diagnostic methods which achieve 88-90% accuracy. This systematic review including 40 selected studies, involving around 20,000 pregnancies, gave much needed insight into the accuracy of application of AI in prenatal diagnosis of congenital heart defects. In various investigations, ensemble neural networks, convolutional architectures (including YOLO variants and DenseNet ), and explainable AI produced sensitivity values between 75% and 100% and specificity between 76% and 100% for defects such as tetralogy of Fallot, Hypoplastic left heart syndrome, atrioventricular septal defect, and ventricular septal defect. In comparisons, one report recorded an AUC of 0.883 for AI versus 0.749 for residents and 0.808 for fellows, while Ensemble methods achieved 95% sensitivity and 96% specificity compared with traditional measurements at 88% and 90%.

Performance was consistently high across gestational ages. First-trimester screenings reached approximately 95.6% accuracy, and studies focused on the common second-trimester period (18–24 weeks) and later gestations reported similar detection levels.

**Conclusion:** These results support that, within their respective designs and sample sizes, AI techniques generally provide detection metrics that meet or exceed those of established diagnostic methods during prenatal CHD screening and in effect provide an augmented intelligence for clinical diagnosis. Further research with more robust standardization and more large scale randomized studies is needed to validate the accuracy and diagnostic applications that can be harnessed.

## Introduction

Congenital heart defects are one of the most common congenital birth defects. Of every 100 children born around the world, one will have a malformation of the heart. Nearly half the children born with a CHD will need a medical intervention in their lifetime, and a quarter of them will need it in the first year of their life in order to survive. (a.constantinof, n.d.) . Congenital heart disease is a large, rapidly emerging global problem in child health. Without the ability to substantially alter the prevalence of congenital heart disease, interventions and resources must be used to improve survival and quality of life. (Meghan.S, n.d.).

Prenatal detection rates remain highly variable, as most CHD occur in low risk pregnancies and therefore depend on the maternal obstetric provider to recognize fetal cardiac abnormality on obstetric screening anatomic ultrasound. Furthermore, rigorous ultrasound training as well as newer advanced imaging technology in ultrasound machines has made it possible to detect major heart defects by the end of the first trimester of pregnancy. However this largely depends on the training and expertise of the physician undertaking the ultrasound scan. Hence diagnostic accuracy is largely experience and operator-dependent. Trained professionals may not always be readily available, and may increase cost and waiting times for key clinical diagnosis.

The greatest impact of fetal echocardiography remains identification of critical CHD before birth to allow immediate cardiac management after delivery to decrease neonatal morbidity and mortality. Analyzing the severity of abnormal cardiac physiology in various forms of CHD before birth allows the fetal cardiologist to prognosticate effects on the developing fetus, predict risk of postnatal hemodynamic instability, guide delivery planning through multidisciplinary collaboration, and anticipate how the disease will impact the neonate after delivery. (Sun, n.d.)

Artificial intelligence (AI) technology, with its rapid development, utilizing advanced computer algorithms, has great potential to empower sonographers in time-saving and accurate diagnosis and to bridge the skill gap in different regions. In recent years, AI-assisted fetal echocardiography has been successfully applied to a wide range of ultrasound diagnoses. (Zhang, n.d.). Furthermore AI can reduce learning timelines and make training or clinical diagnostics available in even remote areas where access to fetal echocardiography is limited, especially due to limited clinical resources and unavailability of trained professionals.

It is estimated that 90% of congenital heart defects occur in otherwise low-risk populations where fetal echocardiogram would never be routinely advised. Chance of detection in bedside scan of certain cardiac defects is as low as 10-26%. AI can bridge this gap of screening as it is used in mainly 3 applications, for image processing, automatic biometric measurements and automated disease diagnosis and prediction, and will aid the clinician in important clinical screening.

## Study Methods

This systematic review adhered to the PRISMA (Preferred Reporting Items for Systematic Reviews and Meta-Analyses) guidelines. Only primary research studies evaluating the use of AI in fetal cardiac defect detection was included. After initial search of database and literature, 496 papers were obtained using the keyword search from PubMed and semantic search tools. After exclusion of duplicate studies, data was extracted using a data extraction table (attached at the end of the paper) and after reviewing the abstracts meticulously by both reviewers independently, relevant studies were screened and shortlisted. 40 studies were selected based on the eligibility criteria.

Inclusion criteria:

Studies that involve prenatal ultrasound screening of human fetuses for congenital heart defects Studies that evaluate AI/machine learning algorithms for detecting congenital heart defects

Studies that compare AI detection methods with traditional diagnostic approaches. Studies must report outcome as at least one quantitative measure of diagnostic accuracy (sensitivity, specificity or accuracy)

Study Design; primary research studies (prospective or retrospective).

Gestational Information: The study must specify the gestational age or gestational age ranges of screened fetuses Diagnostic Timing on prenatal diagnosis studied must be noted.

Include only the studies that use direct fetal cardiac imaging (rather than animal models or simulations only).

Exclusion criteria:

- Non-human studies
- Duplicate publications
- Studies without key information related to the question asked

## Study Design

The specific type of study design used (e.g., prospective cohort, cross-sectional, case-control), was described. We considered all screening questions and made a holistic judgement about what to screen in each paper.

Search: **((((artificial intelligence) AND (fetal)) AND (detection)) AND (congenital heart defect)) AND (malformation)** ("artificial intelligence"[MeSH Terms] OR ("artificial"[All Fields] AND "intelligence"[All Fields]) OR "artificial intelligence"[All Fields]) AND ("fetale"[All Fields] OR "fetally"[All Fields] OR "fetals"[All Fields] OR "fetus"[MeSH Terms] OR "fetus"[All Fields] OR "fetal"[All Fields] OR "foetal"[All Fields]) AND ("detect"[All Fields] OR "detectabilities"[All Fields] OR "detectability"[All Fields] OR "detectable"[All Fields] OR "detectables"[All Fields] OR "detectably"[All Fields] OR "detected"[All Fields] OR "detectible"[All Fields] OR "detecting"[All Fields] OR "detection"[All Fields] OR "detections"[All Fields] OR "detects"[All Fields]) AND ("heart defects, congenital"[MeSH Terms] OR ("heart"[All Fields] AND "defects"[All Fields] AND "congenital"[All Fields]) OR "congenital heart defects"[All Fields] OR ("congenital"[All Fields] AND "heart"[All Fields] AND "defect"[All Fields]) OR "congenital heart defect"[All Fields]) AND ("abnormalities"[MeSH Subheading] OR "abnormalities"[All Fields] OR "malformations"[All Fields] OR "congenital abnormalities"[MeSH Terms] OR ("congenital"[All Fields] AND "abnormalities"[All Fields]) OR "congenital abnormalities"[All Fields] OR "malformation"[All Fields] OR "malformational"[All Fields] OR "malformative"[All Fields] OR "malformed"[All Fields])

### Title and Abstract Screening

The two independent authors conducted the title and abstract screening of the studies retrieved from the systematic search using the predefined eligibility criteria. Any disagreements regarding the inclusion of studies for full-text review were discussed between the authors to reach a consensus.

### Risk of Bias Assessment

The quality of the included studies was assessed using the Cochrane Collaboration’s Risk of Bias 2 (RoB 2) tool.

### Full-Text Screening and Data Extraction

Potentially eligible full-text articles were reviewed independently by the two authors to assess their suitability for inclusion. Data were then extracted from the eligible studies and disagreements at any stage were resolved through discussion. The systematic review, including the literature search, screening, and data extraction, followed the Preferred Reporting Items for Systematic Reviews and Meta-Analyses (PRISMA 2020) guidelines and was illustrated using the PRISMA flowchart.

The data extraction process was documented in a Microsoft Excel spreadsheet, where the following information was collected:

Data Collection and Sample Characteristics: Total number of participants/images were noted, Gestational age range of fetuses at the time of study, time period of data collection, source of data (e.g., retrospective echocardiograms, screening ultrasounds) were recorded. The breakdown of normal vs. abnormal heart conditions, specifically type of abnormality were also noted.

AI Algorithm Characteristics: Description of the specific AI/machine learning approach including type of neural network (e.g., convolutional, ensemble), the type of algorithm used, including key technical features of the algorithm was recorded. Relevant training methodology (e.g., supervised learning) was also studied.

Diagnostic Performance Metrics. We extracted reported diagnostic performance metrics, including sensitivity, specificity, accuracy, positive Predictive Value, negative Predictive Value, Area Under the Curve (AUC) and Confidence Intervals (if provided)

Specific Congenital Heart Defects Detected. Specific congenital heart defects were identified in the study, including name of each defect, prevalence or frequency of detection and any notable characteristics of detection.

Clinical Significance and Comparative Performance: We tried to summarize the study’s key findings about AI performance. Comparison to traditional diagnostic methods. Improvement over existing techniques. Potential clinical implications of the findings and also looked at any limitations or challenges that it may pose.

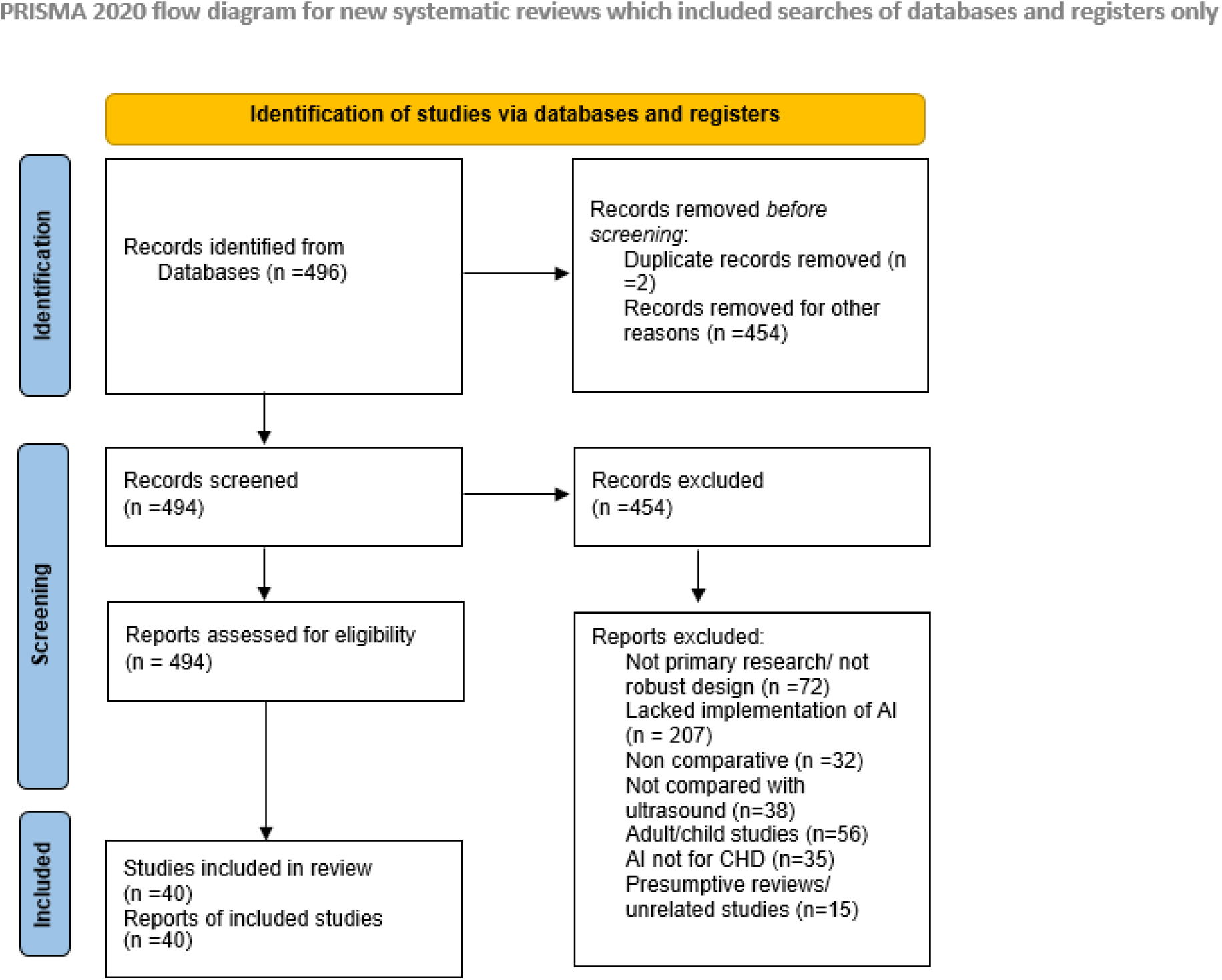

**This study is not registered as not a clinical trial**

**Conflicts of interest/ competing interest:** The authors have no conflict of interest

**Disclosure summary:** The authors have nothing to disclose

**Funding statement:** This research received no external funding

## Ethical Statement

An ethical review was not required for this study as it exclusively used data available from previously published sources

## Results

### Characteristics of Included Studies

| Study | Study Design | AI Algorithm Type | Gestational Age Range | Sample Size |
| --- | --- | --- | --- | --- |
| Arnaout et al., 2018 | Retrospective study | Convolutional and fully convolutional neural networks | 18-24 weeks | 685 |
| Arnaout et al., 2020 | Retrospective study | Ensemble of neural networks (ResNet, U-Net) | 18-24 weeks | 1,326 studies, 107,823 images |
| Arunamata et al., 2023 | Retrospective study | Deep Neural Network | No mention found (2T or 3T) | 117 |
| Athalye et al., 2023b | Retrospective study | Deep learning model | 18 to 22 weeks | 108 ultrasound studies |
| Bokerija et al., 2024 | Prospective study | Deep learning, instance segmentation | 18-21 weeks | No mention found |
| Day et al., 2024 | Prospective randomised controlled trial | Sononet architecture | 18 +0 to 27 +6 weeks | 78 pregnant participants |
| Gong et al., 2020 | Machine learning study | DGACNN | No mention found | No mention found |
| Habib et al., 2024 | Retrospective study | MobileNet | 20-28 weeks | 200 |
| Han et al., 2022 | Prospective study | AI segmentation models | No mention found | 9,657 |
| Huang et al., 2020 | Prospective study | 5D Heart and STICloop | Second and third trimesters | 250 |
| Iliescu et al., 2021 | Prospective study | Deep Learning | 12-13+6 weeks | 7251 images from<br>326 US<br>scans |
| Kim et al., 2021 | Not specified in<br>abstract | HeartAssistTM | 20 to 40 weeks | 4,000 |

| Study | Study Design | AI Algorithm Type | Gestational Age Range | Sample Size |
| --- | --- | --- | --- | --- |
| Komatsu et al., 2021 | Retrospective study | CNN-based SONO architecture | No mention found | No mention found |
| Le et al., 2020 | Prospective study | Machine learning | No mention found | 3910 |
| Lu et al., 2024 | Deep learning study | SKGC framework | No mention found | No mention found |
| Magesh and Raja Kumar, 2023 | Deep learning study | Reg net network | 18 to 34 weeks | 363 |
| Matsuoka et al., 2023 | Not specified in abstract | Explainable AI | 18-34 weeks | 160 cases and 344 videos |
| Nurmaini et al., 2022 | Prospective observational study | DenseNet201 | 18-23 weeks | 9277 women |
| Pietrolucci et al., 2023 | Prospective observational study | Heartassist™ | 19–23 weeks | 120 |
| Qiao et al., 2022 | Machine learning study | SPReCHD network | No mention found | No mention found |
| Rahman et al., 2023 | Prospective study | Stacking CNN-LSTM models | No mention found | 22 |
| Sakai et al., 2022 | Deep learning study | Explainable AI representation | No mention found | No mention found |
| Stos et al., 2024 | Retrospective study | Deep Neural Network | 18–25 weeks | No mention found |
| Su et al., 2021 | Retrospective study | Machine learning | 20–23 + 6 weeks | 1219 cases (219 CHD, 1000 normal) |
| Sushma et al., 2021 | Retrospective study | SVM and deep learning methods | 18-24 weeks | 1628 |
| Sutarno et al., 2022 | Prospective study | Dense convolutional network | No mention found | 460 US images |
| Tang et al., 2022 | Retrospective study | Two-stage deep transfer learning | 20–40 weeks | 6941 pregnancies |
| Ungureanu et al., 2023 | Prospective cross-sectional study | Deep Learning | 12-13 + 6 weeks | No mention found |
| Wang et al., 2024 | Prospective study | Two-stage detection approach | More than 21 weeks | 2676 original source images |
| Weichert and Weichert, 2021 | Not specified in abstract | FINE method | No mention found | No mention found |
| Wu et al., 2022 | Deep learning study | YOLOv5 | No mention found | 1695 |
| Wu et al., 2023 | Retrospective study | Machine learning | 20-24 weeks | 3360 images |
| Yang et al., 2023 | Deep learning study | YOLOv5 | No mention found | No mention found |
| Yeo et al., 2018 | Prospective case-control study | FINE method | 16 to 37 weeks | 150 |
| Zhou et al., 2024a | Retrospective observational study | U-net, MA-net, Link-net | Mean 24.4±4.1 weeks | 275 fetuses (278 datasets) |

Our analysis of 40 studies on Artificial Intelligence (AI) applications in fetal echocardiography revealed:

- Study Design :

**–** Retrospective studies were most common (14 studies)
**–** Prospective studies were the second most common (12 studies)
**–** 7 studies were specifically described as deep learning studies **–** We found 1 randomized controlled trial and 1 systematic review

- AI Algorithm Types :

**–** A wide variety of AI algorithms were used
**–** Deep learning/neural networks were the most common (6 studies)
**–** UNet architectures were frequently used (5 studies)
**–** Other common algorithms included Convolutional Neural Network (CNN) (3 studies), You Only Look Once (YOLO) (3 studies), and machine learning (3 studies)
**–** Many studies used unique or proprietary algorithms

- Gestational Age Range :

**–** We didn’t find gestational age information in the available abstracts or full texts for 19 studies
**–** Where reported, the most common range was 18-24 weeks or within this range (8 studies)
**–** 4 studies included a wider range of 20-40 weeks or within this range
**–** 2 studies focused on early pregnancy (12-13 weeks)
- Sample Size :

**–** 29 studies reported sample size, while we didn’t find this information for 11 studies
**–** The total sample size across all studies that reported it was 169,274
**–** Sample sizes varied widely, from small studies with 22 participants to large studies with over 100,000 images

### Detection Performance

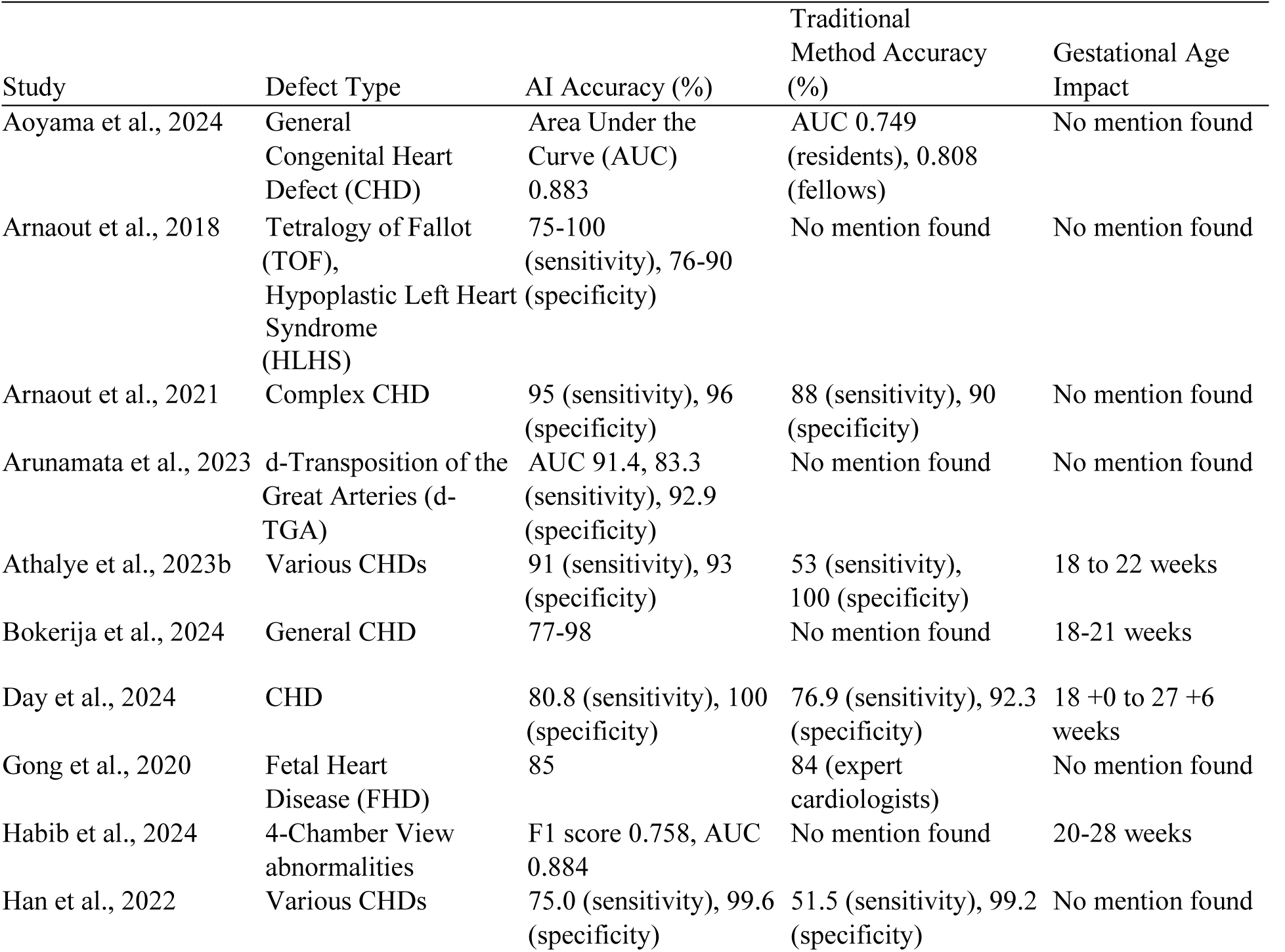
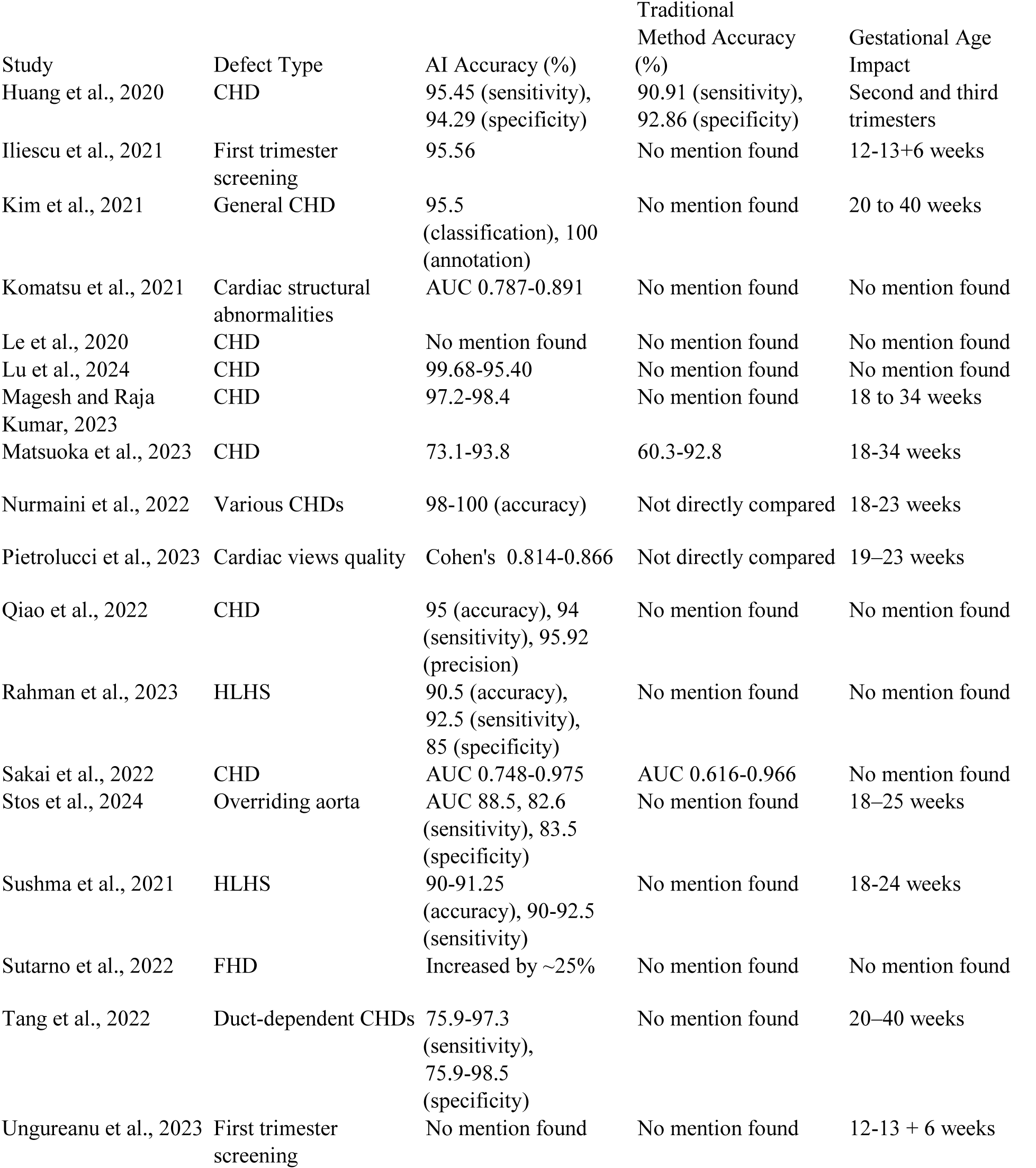
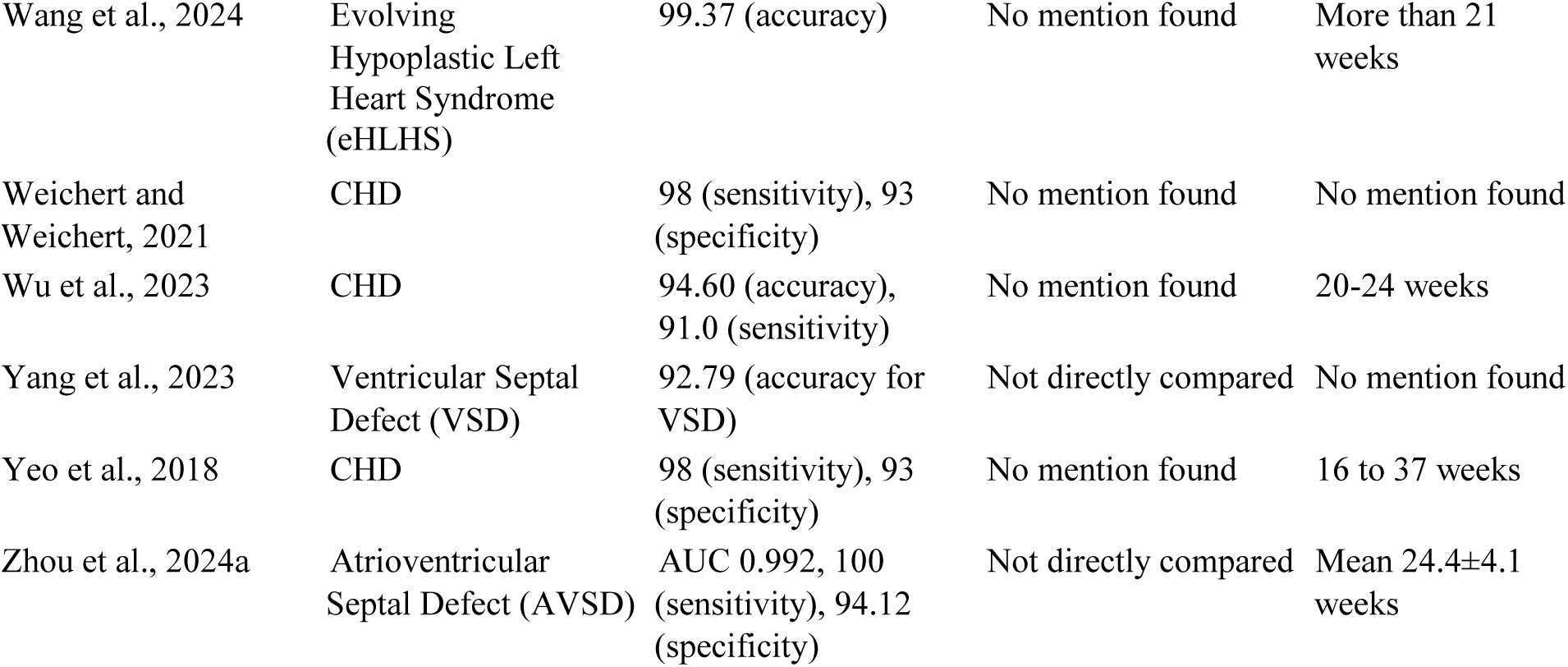

| Study | Defect Type | AI Accuracy (%) | Traditional Method Accuracy (%) | Gestational Age Impact |
| --- | --- | --- | --- | --- |
| Aoyama et al., 2024 | General Congenital Heart Defect (CHD) | Area Under the Curve (AUC) 0.883 | AUC 0.749 (residents), 0.808 (fellows) | No mention found |
| Arnaout et al., 2018 | Tetralogy of Fallot (TOF), Hypoplastic Left Heart Syndrome (HLHS) | 75-100 (sensitivity), 76-90 (specificity) | No mention found | No mention found |
| Arnaout et al., 2021 | Complex CHD | 95 (sensitivity), 96 (specificity) | 88 (sensitivity), 90 (specificity) | No mention found |
| Arunamata et al., 2023 | d-Transposition of the Great Arteries (d-TGA) | AUC 91.4, 83.3 (sensitivity), 92.9 (specificity) | No mention found | No mention found |
| Athalye et al., 2023b | Various CHDs | 91 (sensitivity), 93 (specificity) | 53 (sensitivity), 100 (specificity) | 18 to 22 weeks |
| Bokerija et al., 2024 | General CHD | 77-98 | No mention found | 18-21 weeks |
| Day et al., 2024 | CHD | 80.8 (sensitivity), 100 (specificity) | 76.9 (sensitivity), 92.3 (specificity) | 18 +0 to 27 +6 weeks |
| Gong et al., 2020 | Fetal Heart Disease (FHD) | 85 | 84 (expert cardiologists) | No mention found |
| Habib et al., 2024 | 4-Chamber View abnormalities | F1 score 0.758, AUC 0.884 | No mention found | 20-28 weeks |
| Han et al., 2022 | Various CHDs | 75.0 (sensitivity), 99.6 (specificity) | 51.5 (sensitivity), 99.2 (specificity) | No mention found |
| Huang et al., 2020 | CHD | 95.45 (sensitivity), 94.29 (specificity) | 90.91 (sensitivity), 92.86 (specificity) | Second and third trimesters |
| Iliescu et al., 2021 | First trimester screening | 95.56 | No mention found | 12-13+6 weeks |
| Kim et al., 2021 | General CHD | 95.5 (classification), 100 (annotation) | No mention found | 20 to 40 weeks |
| Komatsu et al., 2021 | Cardiac structural abnormalities | AUC 0.787-0.891 | No mention found | No mention found |
| Le et al., 2020 | CHD | No mention found | No mention found | No mention found |
| Lu et al., 2024 | CHD | 99.68-95.40 | No mention found | No mention found |
| Magesh and Raja Kumar, 2023 | CHD | 97.2-98.4 | No mention found | 18 to 34 weeks |
| Matsuoka et al., 2023 | CHD | 73.1-93.8 | 60.3-92.8 | 18-34 weeks |
| Nurmaini et al., 2022 | Various CHDs | 98-100 (accuracy) | Not directly compared | 18-23 weeks |
| Pietrolucci et al., 2023 | Cardiac views quality | Cohen's 0.814-0.866 | Not directly compared | 19–23 weeks |
| Qiao et al., 2022 | CHD | 95 (accuracy), 94 (sensitivity), 95.92 (precision) | No mention found | No mention found |
| Rahman et al., 2023 | HLHS | 90.5 (accuracy), 92.5 (sensitivity), 85 (specificity) | No mention found | No mention found |
| Sakai et al., 2022 | CHD | AUC 0.748-0.975 | AUC 0.616-0.966 | No mention found |
| Stos et al., 2024 | Overriding aorta | AUC 88.5, 82.6 (sensitivity), 83.5 (specificity) | No mention found | 18–25 weeks |
| Sushma et al., 2021 | HLHS | 90-91.25 (accuracy), 90-92.5 (sensitivity) | No mention found | 18-24 weeks |
| Sutarno et al., 2022 | FHD | Increased by ~25% | No mention found | No mention found |
| Tang et al., 2022 | Duct-dependent CHDs | 75.9-97.3 (sensitivity), 75.9-98.5 (specificity) | No mention found | 20–40 weeks |
| Ungureanu et al., 2023 | First trimester screening | No mention found | No mention found | 12-13 + 6 weeks |
| Wang et al., 2024 | Evolving Hypoplastic Left Heart Syndrome (eHLHS) | 99.37 (accuracy) | No mention found | More than 21 weeks |
| Weichert and Weichert, 2021 | CHD | 98 (sensitivity), 93 (specificity) | No mention found | No mention found |
| Wu et al., 2023 | CHD | 94.60 (accuracy), 91.0 (sensitivity) | No mention found | 20-24 weeks |
| Yang et al., 2023 | Ventricular Septal Defect (VSD) | 92.79 (accuracy for VSD) | Not directly compared | No mention found |
| Yeo et al., 2018 | CHD | 98 (sensitivity), 93 (specificity) | No mention found | 16 to 37 weeks |
| Zhou et al., 2024a | Atrioventricular Septal Defect (AVSD) | AUC 0.992, 100 (sensitivity), 94.12 (specificity) | Not directly compared | Mean 24.4±4.1 weeks |

### Key findings

We found a wide range of congenital heart defects (CHDs) studied across the 40 papers, with general CHD or various CHDs being the most common focus (20 studies). Specific defects like HLHS, AVSD, and FHD were also studied in multiple papers.

AI performance was reported using various metrics:

- 15 studies reported sensitivity and specificity
- 13 studies reported accuracy
- 7 studies reported Area Under the Curve (AUC)
- 3 studies used other measures (F1 score, Cohen’s , precision)
- We didn’t find AI performance metrics in the available abstracts or full texts for 3 studies Traditional method accuracy was less frequently reported:
- 8 studies provided traditional method accuracy
- 25 studies did not report traditional method accuracy
- 7 studies did not directly compare AI to traditional methods

When both AI and traditional method accuracies were reported, AI generally showed higher performance. However, the lack of consistent reporting makes direct comparisons challenging across all studies.

Gestational age was reported in 21 studies, with ranges varying widely from first trimester (12-13 weeks) to late third trimester (up to 40 weeks). We didn’t find gestational age information in the available texts for 19 studies.

### Algorithm-Specific Performance

Different AI architectures show varying levels of performance. Convolutional Neural Networks (CNNs): These form the backbone of many successful approaches. For instance, the SONO architecture based on CNNs achieved AUCs of 0.787-0.891 for detecting cardiac structural abnormalities (Komatsu et al., 2021). Ensemble Methods: Arnaout et al. (2020, 2021) used an ensemble of neural networks including ResNet and U-Net, achieving high sensitivity (95%) and specificity (96%). YOLOv5: Wu et al. (2022) reported 96.5% sensitivity and 85.2% precision using YOLOv5, while Yang et al. (2023) achieved 92.79% accuracy for VSD detection. DenseNet: Nurmaini et al. (2022) used DenseNet201, achieving 98-100% accuracy across various CHDs. Specialized Architectures: Novel approaches like DGACNN (Gong et al., 2020) and SPReCHD (Qiao et al., 2022) show promising results, with accuracies of 85% and 95% respectively. Explainable AI: Sakai et al. (2022) demonstrated that explainable AI approaches can improve AUC from 0.616-0.966 (traditional methods) to 0.748-0.975.

These results suggest that while various AI architectures can achieve high performance, ensemble methods and specialized architectures tailored to fetal echocardiography often yield the best results.

### Gestational Age Stratification

While many studies do not explicitly analyze the impact of gestational age on AI performance, we can observe some trends from the reported data, such as in early Pregnancy Iliescu et al. (2021) achieved 95.56% accuracy for first-trimester screening (12-13+6 weeks), demonstrating the potential of AI even in early pregnancy. In the second trimester most studies focus on this period, typically 18-24 weeks, which aligns with standard anomaly scan timing. High performance is consistently reported, such as Day et al. (2024) achieving 80.8% sensitivity and 100% specificity for CHD detection at 18+0 to 27+6 weeks. Even though fewer studies explicitly report on late pregnancy, but those that do show promising results. Kim et al. (2021) reported 95.5% classification accuracy across a wide range of 20-40 week

Moreover some studies, like Yeo et al. (2018), cover a broad range (16 to 37 weeks) and still achieve high sensitivity (98%) and specificity (93%), suggesting AI can perform well across different gestational ages. However, the lack of systematic comparison across gestational ages within individual studies limits our ability to draw firm conclusions about the impact of fetal development on AI performance.

### Comparative Analysis

#### Accuracy by Defect Type

The accuracy of AI algorithms varies depending on the specific type of congenital heart defect:

General CHD Detection : Most studies report high overall accuracy for detecting CHDs. For instance, Magesh and Raja Kumar (2023) achieved 97.2-98.4% accuracy, while Wang et al. (2024) reported 99.37% accuracy for evolving Hypoplastic Left Heart Syndrome (eHLHS). Detection of Specific CHDs was also studied.Tetralogy of Fallot (TOF) and Hypoplastic Left Heart Syndrome (HLHS): Arnaout et al. (2018) reported 75-100% sensitivity and 76-90% specificity. d-Transposition of the Great Arteries (d-TGA): Arunamata et al. (2023) achieved an AUC of 91.4% with 83.3% sensitivity and 92.9% specificity. Atrioventricular Septal Defect (AVSD): Zhou et al. (2024a,b) reported an impressive AUC of 0.992 with 100% sensitivity and 94.12% specificity

Ventricular Septal Defect (VSD): Yang et al. (2023) achieved 92.79% accuracy specifically for VSD detection. Complex vs. Simple CHDs : While not explicitly compared in most studies, some complex CHDs like HLHS and TOF showed high detection rates in individual studies. Arnaout et al. (2020, 2021) reported 95% sensitivity and 96% specificity for complex CHDs. Meanwhile some studies included less common defects. For instance, Stos et al. (2024) focused on detecting overriding aorta, achieving an AUC of 88.5% with 82.6% sensitivity and 83.5% specificity.

Overall, AI algorithms demonstrate high accuracy across various types of CHDs, with some variation depending on the specific defect.

### Time Efficiency and Resource Requirements

Several studies highlight the potential of AI to improve the efficiency of CHD screening. Reduced Scan Duration: Day et al. (2024) reported that AI-assisted scans were significantly shorter than standard scans (median 11.4 min vs 19.7 min). Faster Image Processing : Yang et al. (2023) noted that their YOLOv5 model could recognize each image in just 0.007 seconds. Workload reduction : Wu et al. (2022) suggested that their method could improve the work efficiency of sonographers and reduce their burden Cognitive Load : Day et al. (2024) found that sonographer cognitive load was significantly lower in the AI-assisted group (median NASA TLX score 35.2 vs 46.5). Resource Optimization : Bokerija et al. (2024) mentioned that AI could help reduce unnecessary studies and healthcare costs. These findings suggest that AI has the potential to significantly improve the efficiency of CHD screening, potentially allowing for more widespread and frequent screening without increasing the burden on healthcare resources.

### Limitations of the Review

This systematic review has limitations that must be acknowledged. The heterogeneity of study designs, including the different kinds of AI algorithms studied may impact the reliability of pooled outcomes. Moreover, bias related to selective reporting may also skew the overall findings.

Future research should prioritize high-quality studies with standardized protocols to enhance the reliability of conclusions drawn.

## Conclusions

Several key factors emerge when considering the clinical implementation of AI for CHD detection, such as integration with existing workflows : Studies like Day et al. (2024) demonstrate the potential for AI to be integrated into existing routine screening processes. Performance Across Expertise Levels were improved: Matsuoka et al. (2023) showed that AI improved performance of experts (AUC from 0.966 to 0.975), fellows (0.829 to 0.890), and residents (0.616 to 0.748), suggesting AI could be particularly beneficial for less experienced or training sonographers, hence reducing the learning timeline. Sakai et al. (2022) introduced an explainable AI tool, thereby potentially increasing trust among medical professionals. Moreover several studies, including Wang et al. (2024), emphasize the potential for real-time diagnosis, which could significantly impact clinical decision-making. Studies like Athalye et al. (2023b) demonstrate that AI models can perform well on community-acquired images, suggesting potential for widespread implementation, and remote application. Perhaps it is time to let AI assist in diagnostic improvement, atleast as a standby tool to accelerate second opinions, or as a guide in day to day practice, which will substantially reduce the learning timeline for junior proffessionals. There is still a long way to go, and many large scale randomised studies on the practical day to day applications are needed before implementation of these resources can be achieved.

Most studies position AI as a supportive tool rather than a replacement for human expertise. For instance, Nurmaini et al. (2022) suggest using AI alongside expert fetal cardiologists to enhance diagnostic accuracy.

Whereas Ungureanu et al. (2023) highlighted the potential of AI to serve as a training tool for sonographers, potentially improving overall screening quality.

These considerations suggest that while AI shows great promise for improving congenital heart defect detection, careful planning and integration will be necessary for successful clinical implementation. The focus should be on using AI to augment and support human expertise rather than replace it entirely.

## Data Availability

All data produced in the present study are available upon reasonable request to the authors

## Supporting information

C:\Users\shabana\Documents\data extraction sheet.xlsx

## Data Availability

All data that was studied is uploaded in the data extraction sheet in supplemental files

## Notes

### Competing Interest Statement

The authors have declared no competing interest.

### Funding Statement

Did not receive any funding

## References

A. Arunamata, Marilyne Levy, B. Stos, E. Askinazi, V. Thorey, and C. Gardella. “Abstract 13945: Evaluation of a Deep Neural Network for Detection of D-Transposition of the Great Arteries on Fetal Echocardiograms.” Circulation, 2023.

Akira Sakai, M. Komatsu, R. Komatsu, R. Matsuoka, S. Yasutomi, A. Dozen, K. Shozu, et al. “Medical Professional Enhancement Using Explainable Artificial Intelligence in Fetal Cardiac Ultrasound Screening.” Biomedicines, 2022.

Anda Ungureanu, A. Marcu, C. Patru, D. Ruican, R. Nagy, R. Stoean, C. Stoean, and D. Iliescu. “Learning Deep Architectures for the Interpretation of First-Trimester Fetal Echocardiography (LIFE) - a Study Protocol for Developing an Automated Intelligent Decision Support System for Early Fetal Echocardiography.” BMC Pregnancy and Childbirth, 2023.

“Application of Neural Networks in Prenatal Diagnosis of Atrioventricular Septal Defect.” Translational Pediatrics, 2024.

B. Stos, M. Levy, E. Askinazi, A. Constensoux, V. Thorey, and C. Gardella. “Overriding Aorta Identification Using Artificial Intelligence Applied to Second-Trimester Fetal Ultrasound.” Archives of Cardiovascular Diseases, 2024.

C. Athalye, A. Nisselrooij, S. Rizvi, M. Haak, A. Moon-Grady, R. Arnaout, and Original Paper.“Deep-learning Model for Prenatal Congenital Heart Disease Screening Generalizes to Community Setting and Outperforms Clinical Detection.” Ultrasound in Obstetrics and Gynecology, 2023.

C. Athalye, A. van Nisselrooij, S. Rizvi, Monique Haak, A. Moon-Grady, and R. Arnaout. “Deep Learning Model for Prenatal Congenital Heart Disease (CHD) Screening Generalizes to the Community Setting and Outperforms Clinical Detection.” medRxiv, 2023.

Chao Huang, Bowen Zhao, H. Pang, Ran Chen, M. Pan, Xiaohui Peng, and Bei Wang. “Study on Diagnostic Performance of Fetal Intelligent Navigation Echocardiography for Congenital Heart Defect,” 2020.

D. Iliescu, R. Nagy, C. Patru, C. Stoean, R. Stoean, and A. Marcu. “OC12.01: First Trimester Heart Screening Supported by Artificial Intelligence.” Ultrasound in Obstetrics and Gynecology, 2021.

E. L. Bokerija, N. E. Yannaeva, A. N. Sencha, P. V. Metelkin, and O. V. Yurchenko. “Artificial Intelligence in Fetal Echocardiography.” Innovative Medicine of Kuban, 2024.

F. Su, Q. Wu, J. Han, X. Zhang, and D. Kong. “OC10.03: *Application of Computer-aided Diagnosis of Congenital Heart Disease in Four Chamber View of Fetal Heart Basic Screening.” Ultrasound in Obstetrics and Gynecology, 2021.

Gang Wang, Weisheng Li, Mingliang Zhou, Haobo Zhu, Guang Yang, and C. Yap. “4D Foetal Cardiac Ultrasound Image Detection Based on Deep Learning with Weakly Supervised Localisation for Rapid Diagnosis of Evolving Hypoplastic Left Heart Syndrome.” CAAI Transactions on Intelligence Technology, 2024.

Guowei Han, Tianliang Jin, Li Zhang, Chen Guo, Hua Gui, Risu Na, Xuesong Wang, and Haihua Bai. “Adoption of Compound Echocardiography Under Artificial Intelligence Algorithm in Fetal Congenial Heart Disease Screening During Gestation.” Applied Bionics and Biomechanics, 2022.

Huiling Wu, Bingzheng Wu, Fangping Lai, Peizhong Liu, G. Lyu, S. He, and Jiangfeng Dai. “Application of Artificial Intelligence in Anatomical Structure Recognition of Standard Section of Fetal Heart.” Computational and Mathematical Methods in Medicine, 2023.

Huiling Wu, Bingzheng Wu, S. He, and Peizhong Liu. “Congenital Heart Defect Recognition Model Based on YOLOV5.” *International Conference on Anti-Counterfeiting*, Security, and Identification, 2022.

J. Weichert, and A. Weichert. “A ‘Holistic’ Sonographic View on Congenital Heart Disease – How Automatic Reconstruction Using Fetal Intelligent Navigation Echocardiography (FINE) Eases the Unveiling of Abnormal Cardiac Anatomy Part I: Right Heart Anomalies.” Echocardiography, 2021.

Jiajie Tang, Yongen Liang, Yuxuan Jiang, Jinrong Liu, Rui Zhang, Danping Huang, C. Pang, et al. “Screening for Duct-Dependent Congenital Heart Disease from Fetal Echocardiography with Two-Stage Deep Transfer Learning Model: A Multicenter, Retrospective Study.” Social Science Research Network, 2022.

L. Yeo, S. Luewan, and R. Romero. “Fetal Intelligent Navigation Echocardiography (FINE) Detects 98% of Congenital Heart Disease.” Journal of Ultrasound in Medicine, 2018.

M. Komatsu, Akira Sakai, R. Komatsu, R. Matsuoka, S. Yasutomi, K. Shozu, A. Dozen, et al. “Detection of Cardiac Structural Abnormalities in Fetal Ultrasound Videos Using Deep Learning.” Applied Sciences, 2021.

M. Pietrolucci, P. Maqina, I. Mappa, M. C. Marra, Francesco D’ Antonio, and G. Rizzo. “Evaluation of an Artificial Intelligent Algorithm (Heartassist™) to Automatically Assess the Quality of Second Trimester Cardiac Views: A Prospective Study.” Journal of Perinatal Medicine, 2023.

P. Habib, Reza Shirkavand, Samantha Selhorst, O. Turan, Heng Huang, and Sifa Turan. “95 Application of Machine Learning in the Detection of Four Chamber Abnormalities.” American Journal of Obstetrics and Gynecology, 2024.

R. Aoyama, M. Komatsu, Naoaki Harada, R. Komatsu, Akira Sakai, Katsuji Takeda, Naoki Teraya, et al. “Automated Assessment of the Pulmonary Artery-to-Ascending Aorta Ratio in Fetal Cardiac Ultrasound Screening Using Artificial Intelligence.” Bioengineering, 2024.

R. Kim, M. Lee, and H. Won. “OP05.03: HeartAssist TM : A Novel Technique of Automatic Classification and Measurement for Fetal Heart.” Ultrasound in Obstetrics and Gynecology, 2021.

R. Matsuoka, A. Sakai, M. Komatsu, R. Komatsu, R. Aoyama, N. Harada, H. Setoh, K. Iwamoto, A. Sek-izawa, and R. Hamamoto. “EP24.14: Explainable AI to Support Examiners for Abnormality Detection in Fetal Cardiac Ultrasound Screening.” Ultrasound in Obstetrics & Gynecology, 2023.

Rima Arnaout, L. Curran, Erin Chinn, Yili Zhao, and A. Moon-Grady. “Deep-Learning Models Improve on Community-Level Diagnosis for Common Congenital Heart Disease Lesions.” arXiv.org, 2018.

Rima Arnaout, L. Curran, Yili Zhao, J. Levine, Erin Chinn, and A. Moon-Grady. “An Ensemble of Neural Networks Provides Expert-Level Prenatal Detection of Complex Congenital Heart Disease.” Nature Network Boston, 2021.

Rima Arnaout, L. Curran, Yili Zhao, J. Levine, Erin Chinn, and A. Moon-Grady. “Expert-Level Prenatal Detection of Complex Congenital Heart Disease from Screening Ultrasound Using Deep Learning.” medRxiv, 2020.

S. Magesh, and P.S. Raja Kumar. “Fetal Heart Disease Detection Via Deep Reg Network Based on Ultrasound Images.” Journal of Applied Engineering and Technological Science (JAETS), 2023.

S. Nurmaini, M. N. Rachmatullah, Ade Iriani Sapitri, Annisa Darmawahyuni, B. Tutuko, Firdaus Firdaus, R. U. Partan, and Nuswil Bernolian. “Deep Learning-Based Computer-Aided Fetal Echocardiography: Application to Heart Standard View Segmentation for Congenital Heart Defects Detection.” Italian National Conference on Sensors, 2021.

S. Nurmaini, R. U. Partan, Nuswil Bernolian, Ade Iriani Sapitri, B. Tutuko, M. N. Rachmatullah, Annisa Darmawahyuni, Firdaus Firdaus, and J. Mose. “Deep Learning for Improving the Effectiveness of Routine Prenatal Screening for Major Congenital Heart Diseases.” Journal of Clinical Medicine, 2022.

S. Sutarno, S. Nurmaini, R. U. Partan, Ade Iriani Sapitri, B. Tutuko, Muhammad Naufal Rachmatullah, Annisa Darmawahyuni, Firdaus Firdaus, Nuswil Bernolian, and Deny Sulistiyo. “FetalNet: Low-Light Fetal Echocardiography Enhancement and Dense Convolutional Network Classifier for Improving Heart Defect Prediction.” Informatics in Medicine Unlocked, 2022.

Sibo Qiao, Shanchen Pang, Yi Sun, G. Luo, Wenjing Yin, Yawu Zhao, S. Pan, and Zhihan Lv. “SPReCHD: Four-Chamber Semantic Parsing Network for Recognizing Fetal Congenital Heart Disease in Medical Metaverse.” IEEE Journal of Biomedical and Health Informatics, 2022.

T. Day, J. Matthew, S. Budd, A. Farruggia, L. Venturini, R. Wright, B. Jamshidi, et al. “Artificial Intelligence to Assist in the Screening Fetal Anomaly Ultrasound Scan (PROMETHEUS): A Randomised Controlled Trial.” medRxiv, 2024.

T. K. Le, V. Truong, T. Nguyen-Vo, Binh P. Nguyen, T. Ngo, Q. Bui, T. Pham, et al. “APPLICATION OF MACHINE LEARNING IN SCREENING OF CONGENITAL HEART DISEASES USING FETAL ECHOCARDIOGRAPHY.” Journal of the American College of Cardiology, 2020.

T. Sushma, N. Sriraam, P. M. Arakeri, and S. Suresh. “Classification of Fetal Heart Ultrasound Images for the Detection of CHD,” 2021.

Tawsifur Rahman, M. K. A. Al-Ruweidi, Md. Shaheenur Islam Sumon, R. Kamal, M. Chowdhury, and Huseyin C. Yalcin. “Deep Learning Technique for Congenital Heart Disease Detection Using StackingBased CNN-LSTM Models From Fetal Echocardiogram: A Pilot Study.” IEEE Access, 2023.

Xiaoxue Zhou, Tingyang Yang, Yanping Ruan, Ye Zhang, Xiaowei Liu, Ying Zhao, Xiaoyan Gu, Xinxin Xu, Jiancheng Han, and Yihua He. “Application of Neural Networks in Prenatal Diagnosis of Atrioventricular Septal Defect.” Translational Pediatrics, 2024.

Yiru Yang, Bingzheng Wu, Huiling Wu, Wu Xu, G. Lyu, Peizhong Liu, and S. He. “Classification of Normal and Abnormal Fetal Heart Ultrasound Images and Identification of Ventricular Septal Defects Based on Deep Learning.” Journal of Perinatal Medicine, 2023.

Yuhuan Lu, Guanghua Tan, Bin Pu, Hang Wang, Bocheng Liang, Kenli Li, and Jagath C. Rajapakse. “SKGC: A General Semantic-Level Knowledge Guided Classification Framework for Fetal Congenital Heart Disease.” IEEE Journal of Biomedical and Health Informatics, 2024.

Yuxin Gong, Yingying Zhang, Haogang Zhu, J. Lv, Qian Cheng, Hongjia Zhang, Yihua He, and Shuliang Wang. “Fetal Congenital Heart Disease Echocardiogram Screening Based on DGACNN: Adversarial OneClass Classification Combined with Video Transfer Learning.” IEEE Transactions on Medical Imaging, 2020.

(https://world-heart-federation.org/news/call-to-action-on-addressing-the-global-burden-of-pediatric-and-congenital-heart-diseases, n.d.) Addressing the Global Burden of Pediatric and Congenital Heart Diseases Global, regional, and national burden of congenital heart disease, 1990–2017: a systematic analysis for the Global Burden of Disease Study 2017 - The Lancet Child & Adolescent Health

https://pmc.ncbi.nlm.nih.gov/articles/PMC8429868/

https://www.sciencedirect.com/science/article/pii/S0894731724000026

